# Early Structural Cardiovascular Disease, HIV, and Tuberculosis in East Africa (ASANTE): Cross-sectional study protocol for a multimodal cardiac imaging study in Nairobi, Kenya

**DOI:** 10.1101/2025.03.16.25323832

**Authors:** Saate S Shakil, Sidney Korir, Geoffrey Omondi, Boni Maxime Ale, Bernard Gitura, Marian Morris Tofeles, John Kinuthia, Bhavna Chohan, Norrisa Haynes, Carey Farquhar, Priscilla Y Hsue, Chris T Longenecker, Alfred O Osoti

## Abstract

**Introduction:** Persons living with HIV (PLWH) have an augmented risk of cardiovascular disease, including atherosclerosis and myocardial dysfunction, despite effective viral suppression with antiretroviral therapy (ART). Despite the majority of PLWH residing in sub-Saharan Africa, there are limited reports from the region on structural cardiovascular changes due to this residual risk.

Methods and analysis

The Early Structural Cardiovascular Disease, HIV, and Tuberculosis (ASANTE) cross-sectional study will be conducted in a public hospital in Nairobi, Kenya. It will enroll 400 participants (50% female, 50% PLWH) to undergo comprehensive cardiovascular phenotyping using multimodal imaging (coronary CT angiography [CCTA], echocardiography) and banking of biological samples (whole blood, peripheral blood mononuclear cells, serum, and urine). We will define the prevalence of subclinical coronary atherosclerosis by coronary CT angiography (CCTA) and subclinical myocardial dysfunction by transthoracic echocardiography, and evaluate both traditional and non-traditional risk factors, including endemic infections such as latent tuberculosis. This study will contribute important data on phenotypes of and risk factors for HIV-associated cardiovascular disease in this under-studied region.

Ethics and dissemination

Ethical approval for the ASANTE study was granted by the University of Nairobi-Kenyatta National Hospital Ethical Review Committee, Nairobi, Kenya. Results will be submitted for publication in peer-reviewed journals.

**Key messages:** *What is already known on this topic:*

- In studies from high-income countries, HIV infection is an independent risk factor for structural cardiovascular abnormalities (atherosclerosis, myocardial remodeling) and corresponding incident cardiovascular events.
- Emerging data from sub-Saharan Africa suggest there may be a unique epidemiological profile, in which HIV and traditional cardiometabolic risk factors (e.g., hypertension, diabetes, overweight/obesity, smoking) are not associated with coronary atherosclerosis.
- Unique endemic risk factors for cardiovascular disease in HIV have yet to be defined in sub-Saharan Africa.

*What this study adds:*

- We will define the prevalence of and risk factors for subclinical coronary atherosclerosis and myocardial disease in a cohort of Kenyan adults with and without HIV who have been enriched for cardiometabolic risk factors using multimodal imaging (coronary CT angiography, speckle tracking echocardiography).
- We will assess the contribution of latent tuberculosis infection as a potential endemic risk factor for subclinical cardiovascular abnormalities identified on multimodal imaging.

*How this study might affect research, practice, or policy:*

- Delineating the burden, phenotypes, and unique risk factors for early cardiovascular disease in this setting will facilitate tailored interventions for screening, monitoring, and management among persons with HIV.
- Developing multimodal cardiac imaging infrastructure in this resource-limited setting will expand tools for understanding the natural history of cardiovascular disease among persons with HIV in this unique setting.
- This study will yield a diverse collection of stored biological samples, including peripheral blood mononuclear cells, plasma, whole blood RNA, and urine. In combination with the cardiac imaging repository, this biobank will facilitate future collaborative efforts to identify mechanisms underlying phenotypes of cardiovascular disease in this population.

## Introduction

The global burden of HIV-associated cardiovascular disease (CVD) has tripled over the last 2 decades, with the largest reported increase in low- and middle-income regions such as sub-Saharan Africa (SSA).^1^ Despite this, the majority of research on HIV-associated CVD phenotypes is performed in high-income settings.^2^ As nearly two-thirds of persons living with HIV (PLWH) reside in SSA,^3^ there remain significant gaps in understanding the prevalence, phenotypes, and mechanisms of HIV-associated CVD in diverse global populations. Moreover, the risk factor profile for CVD among persons living with HIV (PLWH) in SSA may well be unique due to distinct environmental and endemic exposures.^4,5^

The residual risk of CVD despite effective HIV suppression is thought to be driven by a persistent state of inflammation and immune activation.^6–9^ Current evidence suggests that this inflammation independently drives the two major phenotypes of HIV-associated CVD: accelerated atherosclerotic CVD in all anatomic distributions (coronary, cerebrovascular, peripheral arterial),^10–13^ and myocardial disease leading to adverse ventricular remodeling and heart failure.^14–17^ Over 90% percent of coronary atherosclerosis, present subclinically in up to 50% of low- to moderate-risk adults without HIV, is attributed to established cardiometabolic risk factors (hypertension, diabetes, dyslipidemia, and excess weight/obesity).^18–22^ Much of the data on coronary atherosclerosis among PLWH, however, is derived from high-income country populations, demonstrating that HIV is an independent risk factor for coronary disease, even after adjusting for traditional cardiometabolic risk factors. By contrast, two recent studies among people with and without HIV in Uganda demonstrate a markedly low prevalence of coronary atherosclerosis. In the first, which was enriched for cardiometabolic risk factors, the prevalence was 17% and neither HIV nor traditional risks were associated with coronary atherosclerosis. In the second, the prevalence of coronary atherosclerosis was only 7.7% among rural Ugandans at low to moderate cardiovascular risk.^5,23,24^ Similarly, another study demonstrated that HIV is not associated with carotid atherosclerosis progression in rural Ugandans.^25^ Finally, in the multinational REPRIEVE trial of primary ASCVD prevention in PLWH, only 12 myocardial infarctions were observed among over 1000 enrolled Black African participants over 5 years of follow-up.^26,27^ These findings challenge current paradigms of HIV-associated CVD phenotypes, and call for further anatomical studies in SSA to evaluate the prevalence of and unique risk factors for atherosclerosis, and especially coronary atherosclerosis.

In contrast to the geographic divergence in coronary atherosclerosis epidemiology, HIV infection appears to exert common inflammatory, infiltrative, steatotic, and fibrotic effects on the myocardium in both high-^14–16^ and low-income settings,^28,29^ leading to adverse ventricular remodeling, systolic dysfunction, and diastolic dysfunction.^17,29^ Many of these abnormalities occur in asymptomatic individuals and independently predict incident cardiovascular events;^30^ as such, sensitive markers are needed to assess these subclinical myocardial abnormalities. Speckle-tracking echocardiography, or strain, quantifies myocardial deformation and can detect subclinical dysfunction due to any etiology that precedes the development of overt abnormalities, including reduced left ventricular ejection fraction (LVEF) or impaired diastolic parameters.^31^ Global longitudinal strain (GLS) in particular, a sensitive measure of systolic function, has been shown to be abnormal among PLWH compared with controls,^29,32^ with a prevalence of impaired GLS as high as 50% in HIV.^33^ Impaired GLS is associated with traditional cardiometabolic risk factors,^34^ as well as an adverse risk factor trajectory,^35^ in the general population. Thus, strain could delineate the relative contribution of traditional and non-traditional risk factors to early myocardial damage among PLWH in SSA. Alongside standard measures such as left ventricular hypertrophy (LVH) and the presence and degree of diastolic dysfunction (DD), strain may identify those individuals at high risk of developing subsequent clinical heart failure.

In addition to the role of HIV and traditional risk factors in CVD risk in SSA, unique endemic exposures may contribute to or modify cardiovascular risk in this region. In the REPRIEVE cohort of primary CVD prevention among PLWH, US-based individuals who had coronary disease at baseline demonstrated higher levels of immune activation independently of traditional risk factors.^8^ In developing settings like SSA, this risk may be further augmented by endemic infections such as tuberculosis (TB), a leading cause of death among PLWH.^36^ In the general population, active TB has been linked to ASCVD events despite TB treatment.^37–39^ Moreover, while latent TB infection (LTBI), prevalent in nearly 50% of the population in SSA,^40–42^ has historically been believed to be a state of mycobacterial dormancy, emerging data indicate that LTBI confers a low-level inflammatory state characterized by increased circulating type II interferon in the general population and enhanced monocyte activation among PLWH.^43,44^ Further, LTBI has recently been associated with elevated levels of both pro- and anti-inflammatory cytokines among treated PLWH in Kenya.^45^ While it has been postulated that similar mechanisms may underlie the observed association of LTBI with CAD, ^46^ the effect of LTBI on left ventricular remodeling and subclinical dysfunction among PLWH remain to be investigated.

The *Early Structural Cardiovascular Disease, HIV, and Tuberculosis (ASANTE)* study seeks to address these knowledge gaps using multimodal cardiac imaging in a cross-sectional study of Kenyan adults with and without HIV, and to assess how these phenotypes and mechanisms may interact with co-occurring endemic infections. Our central hypothesis is that traditional cardiometabolic risk factors (hypertension, diabetes, dyslipidemia, smoking, overweight/obesity) and HIV will be associated with subclinical myocardial disease, but not with coronary atherosclerosis. Further, we hypothesize that LTBI will be associated with both atherosclerosis and with myocardial abnormalities, and this relationship is stronger among PLWH. We aim:

1. To determine the prevalence of and risk factors for subclinical coronary atherosclerosis, as assessed by coronary CT angiography (CCTA), among Kenyan adults with HIV and age/sex-matched HIV-uninfected adults.
2. To define the prevalence of and risk factors for subclinical myocardial disease, as defined by sensitive echocardiographic measures (peak systolic global longitudinal strain [GLS], left ventricular hypertrophy [LVH], and diastolic dysfunction [DD]) among Kenyan adults with HIV and age/sex-matched HIV-uninfected adults.
3. To define the relationship of latent tuberculosis infection (LTBI) with subclinical coronary and myocardial abnormalities and assess how these associations are modified by HIV co-infection.

## Methods and analysis

### Study design

This is a cross-sectional study evaluating prevalence, phenotypes, and risk factors for cardiovascular disease in Kenya. Data collection commenced in December 2023 and will continue for approximately 36 months. Using a cross-sectional approach, we will define the prevalence of subclinical cardiovascular disease using multimodality imaging in a mixed cohort of Kenyan adults with HIV (PLWH; n = 200) and HIV-uninfected controls (n = 200).

### Setting

This study is being conducted at Kenyatta National Referral and Teaching Hospital (KNH) in Nairobi, Nairobi County, Kenya. KNH is the largest government referral hospital in Kenya and serves a diverse urban and peri-urban population from within and around Nairobi, making it an ideal facility for recruiting participants of various demographic backgrounds. PLWH are recruited from the KNH Comprehensive Care Center (CCC), which provides care for approximately 22,000 PLWH, with an average daily attendance of 200 patients. Control participants are recruited from the KNH Voluntary HIV Testing Centers, the Accident & Emergency, diabetes and hypertension specialty clinics, and adult general medicine clinics, all of which see a similarly high volume of patients.

### Study population

The study is enrolling 200 PLWH (100 females, 100 males) and 200 age/sex-matched controls who have tested negative for HIV. Inclusion criteria are as follows: age ≥45 years at enrollment; presence of at least 1 cardiometabolic risk factor (hypertension, diabetes, dyslipidemia, overweight/obese, smoking). PLWH must have been on stable antiretroviral therapy for ≥6 months. Eligible control participants must test HIV-antibody negative at enrollment. Individuals are excluded if they have known history of prior cardiovascular disease (e.g., myocardial infarction, heart failure, other known clinical/structural cardiac abnormality); pregnant; meet any criteria that would preclude contrast administration for CCTA (**Table 1**), or atrial fibrillation/flutter on screening electrocardiogram. Eligible participants are enrolled 2:2:1 across three age strata (45-54, 55-64, and 65+ years) and balanced by biological sex. Inclusion and exclusion criteria are summarized in **Table 1**.

**Table 1.** Study inclusion/exclusion criteria.

|  |  |
| --- | --- |
| **Inclusion criteria** | · Age $\geq 45$ years<br>· At least one cardiometabolic risk factor (e.g., hypertension, diabetes, dyslipidemia, overweight/obesity, smoking)<br>· PLWH must have been on antiretroviral therapy for $\geq 6$ months |
| **Exclusion criteria** | · Known history of cardiovascular disease (e.g., heart failure, prior myocardial infarction) or atrial fibrillation/flutter<br>· Atrial fibrillation/flutter on screening electrocardiogram<br>· Pregnancy<br>· eGFR $< 60$ mL/min/1.73 m<sup>2</sup> based on point-of-care serum creatinine<br>· History of iodinated contrast allergy, intolerance to beta blockers or nitroglycerin (SBP $< 90$ mmHg, severe reactive airway disease, use of phosphodiesterase-5 inhibitors $< 48$ hours)<br>· History of hyperthyroidism, pheochromocytoma, claustrophobia, or weight/girth exceeding CT scanner limit |

### Data collection

All study assessments are performed within 1 week of enrollment. **Figure 1** illustrates a study schematic mapping the process from screening to data collection.

**Figure 1.**
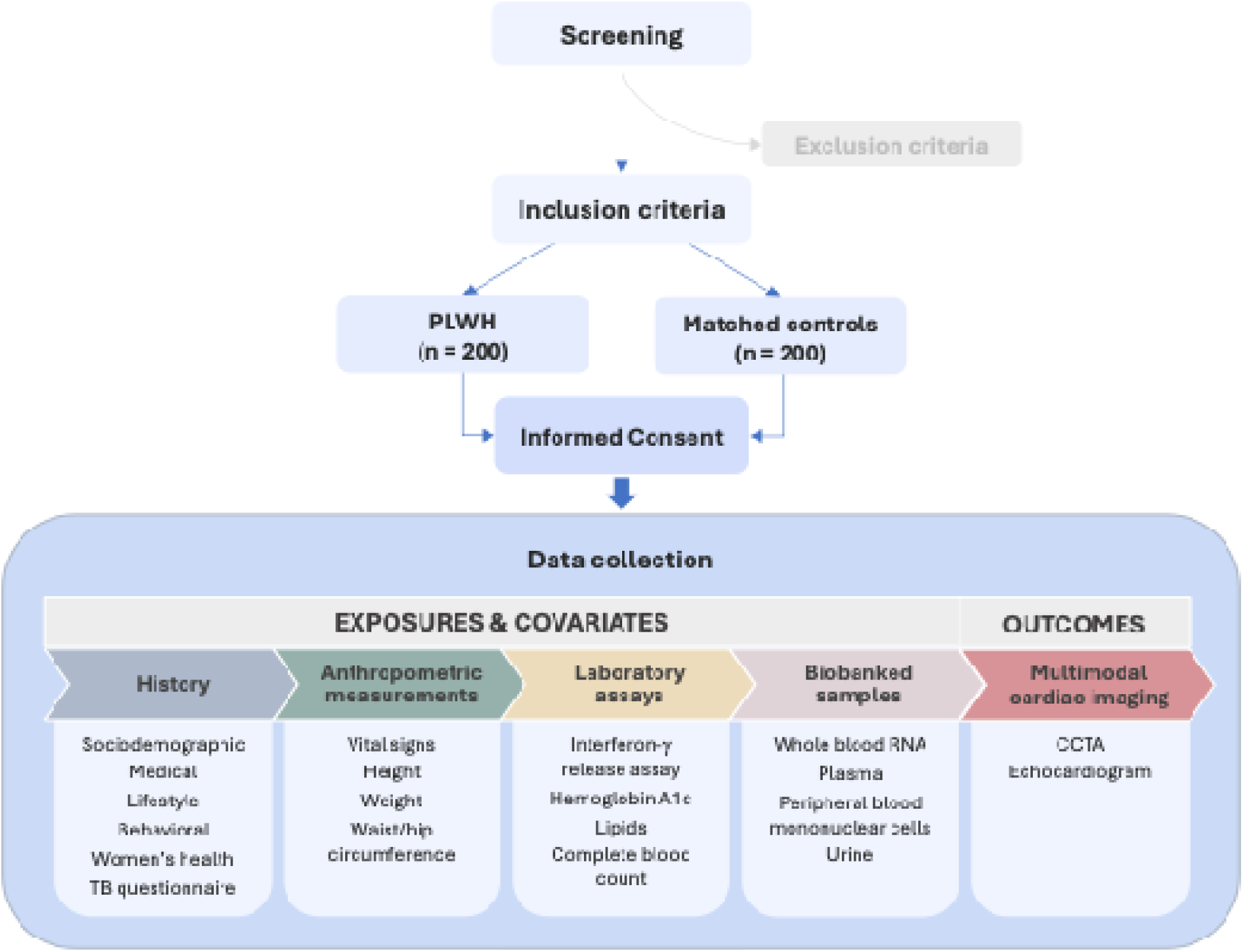
Study outline and data collection workflow (PLWH, persons living with HIV; TB, tuberculosis; CCTA, coronary CT angiography).

#### History

A comprehensive demographic, health, and lifestyle questionnaire will be administered to all participants. The questionnaire includes basic demographic information such as age, education, wealth/income measures; medical and mental health history such as presence of cardiovascular risk factors (e.g., hypertension, diabetes), infectious exposure (e.g., tuberculosis symptom questionnaire and treatment history), depression symptoms (PHQ-8); lifestyle details such as smoking, physical activity, and sleep history. Female participants will also complete a women’s health history. For PLWH, HIV-specific data are abstracted from the medical record (e.g., date of diagnosis, duration of antiretroviral therapy, antiretroviral regimen).

#### Anthropometric measures

Participants will undergo a detailed physical assessment at enrollment, including temperature, heart rate, respiratory rate, pulse oximetry, and blood pressure. Standardized biometric measurements of height, weight, waist, and hip circumference are also obtained.

#### Biological samples

Peripheral venous blood samples (approximately 30 mL total) will be collected from each eligible participant at enrollment to assay creatinine, hemoglobin A1c, and complete blood count. Additional samples will be collected and stored for future assays, including isolated and preserved peripheral blood mononuclear cells (stored in liquid nitrogen at −80°C); preserved plasma; preserved whole blood RNA (in PAXgene RNA tubes, Qiagen); and preserved urine at −70°C.

#### Ascertainment of latent tuberculosis infection (LTBI) status

LTBI status will be assessed using interferon-γ release assay (QuantiFERON-TB Gold Plus ELISA, Qiagen) performed on blood samples at enrollment.

#### Echocardiography

Participants will undergo a standardized comprehensive transthoracic echocardiogram according to the American Society of Echocardiography guidelines^47^ using a Philips CX50 ultrasound machine. Images will be acquired by a trained study sonographer, who is blinded as to the participant’s HIV serostatus and medical history. All images will be stored in DICOM format, uploaded to a secure cloud, and archived locally on a secure hard disk. Measurements of global longitudinal strain, LV mass, and assessments of diastolic function, and other standard echocardiographic assessments will be performed offline by a blinded reader using the latest version of TOMTEC Imaging Systems software. Primary and secondary echocardiographic outcomes are summarized in **Table 2**.

**Table 2.** Outcomes for noninvasive imaging studies.

| Outcomes | CT | Echo |
| --- | --- | --- |
| **Primary** | · Presence of plaque (binary) | · Left ventricular global longitudinal strain (GLS; continuous)<br>· Presence of diastolic dysfunction (binary)<br>· LV hypertrophy<sup>§</sup> (LVH; binary) |
| **Secondary** | · Segment involvement score (SIS)*<br>· Segment severity score (SSS)<sup>†</sup><br>· Plaque characteristics (degree of calcification, high-risk features<sup>‡</sup>) | · Diastolic dysfunction grade<br>· LV geometric patterns (normal, concentric, eccentric)<br>· LV mass index |
| **Exploratory** | · LV mass index<br>· Pulmonary artery size<br>· Pericardial fat volume | · Left atrial strain<br>· LV diastolic strain<br>· Right ventricular strain |

<sup>§</sup>LVH will be defined by sex-specific cutoffs for indexed LV mass; \*SIS: total number of diseased segments; <sup>†</sup>SSS: calculated using a luminal obstruction weight for that segment (x1 if $< 25\%$ obstruction, x2 if 25-50%, x3 if 50-70%, x4 if 70-99% and x5 if completely occluded), with maximum possible SSS of 90 for an 18-segment model; <sup>‡</sup>High-risk plaques are defined as showing any of the following features: positive remodeling, spotty calcifications, or napkin-ring sign.

#### Coronary CT angiography (CCTA)

The ASANTE study will employ noninvasive coronary CT angiography (CCTA), a contrast-based evaluation of the coronary arteries whose sensitivity is comparable to the gold standard of invasive angiography for the evaluation of plaque.^48^ Participants will undergo retrospective ECG-gated CCTA on a 128-slice Siemens SOMATOM Definition scanner based on protocols in accordance with the Society of Cardiovascular CT Guidelines.^49^ Metoprolol will be administered prior to CCTA to achieve a goal heart rate of <70 bpm, which optimizes image quality by allowing maximum diastolic filling of the coronary arteries. Following an initial non-contrast coronary artery calcium scan (30 mAs, 130 kV, and 60% R-R gating), 0.4 mg sublingual nitroglycerin will be administered one minute prior to the contrast scan (80 mL Ultravist/iopromide) to maximally vasodilate the coronaries and thereby allow optimal contrast opacification of the vessels. Following a 5-second breath hold upon automatic triggering, a bolus-triggered CCTA will be obtained (400 mAs, 130 kV, and ECG pulsing at 30-70% R-R peak) with slices through a 35 cm field of view from the bifurcation of the pulmonary arteries to the apex of the heart. Reconstructions will be at I31s medium smooth kernel and sinogram affirmed iterative reconstruction (SAFIRE) level 3, with 0.75 mm slices at 0.5 mm increments. CT scans will be read by a local radiologist for clinically significant findings, then uploaded to a secure cloud storage system to be read centrally for research by a blinded core laboratory with extensive CCTA expertise. Primary and secondary CT outcomes are summarized in **Table 2**.

### Outcome measures

#### CCTA

As coronary atherosclerosis is relatively less common in the East African population compared with high-income regions,^23^ the primary CCTA outcome will be presence of any coronary plaque (binary variable; **Table 2**). Secondary CCTA outcomes include coronary segment involvement and severity score, which are semiquantitative measures of the extent of overall plaque burden and stenosis severity, respectively. Exploratory outcomes include LV mass index, pulmonary artery size, and pericardial fat volume.

#### Echocardiography

The primary echocardiographic outcomes will be peak systolic global longitudinal strain (GLS; continuous), presence of left ventricular hypertrophy (LVH; binary), and presence of diastolic dysfunction (DD; binary). Secondary outcomes include degree of DD (ordinal); left ventricular geometric patterns (categorical; normal, concentric, or eccentric remodeling); and left ventricular mass index (continuous). Exploratory outcomes to be evaluated will include left atrial strain, left ventricular diastolic strain, right ventricular strain, and pulmonary artery systolic pressures.

### Sample size calculation

Studies performed in high-income settings report an approximately 50% higher incidence of myocardial infarction and prevalence of coronary atherosclerosis among PLWH.^50,51^ By contrast, though data on coronary atherosclerosis in sub-Saharan Africa are limited, a recent study in Uganda reports prevalence of 14% and 26% among PLWH and controls, respectively.^52^ Based on a two-sided α of 0.05, our sample of 200 participants per group will allow us to detect similar differences in coronary atherosclerosis with 85% power.

Although studies consistently demonstrate depressed peak systolic global longitudinal strain (GLS) in PLWH compared with controls, they are limited in number with small sample sizes.^53^ Similarly, other studies in the region report a prevalence of abnormal GLS up to 50% in PLWH,^54^ but there are few such reports in HIV-uninfected groups. We estimate that based on an α of 0.05, we will be able to detect a difference in mean GLS of −1 to −1.5% with a sample size of 200 individuals per group based on standard deviation estimates between 3 and 6, at a power of 80%. These estimates should be considered conservative approximations to detect a clinically relevant difference in GLS by HIV serostatus. Prior studies have reported a prevalence of left ventricular diastolic dysfunction in PLWH vs. uninfected adults as 49% vs 29% in the United States^55^ and 45% vs 37% in East Africa,^56^ respectively, corresponding to odds ratios of 1.5 to 2.2. Based on a two-sided α of 0.05, our sample of n = 400 (50% PLWH) will allow us to detect similar differences in diastolic dysfunction prevalence with 75-86% power. Few studies have reported on prevalence of LVH in SSA, and estimates in the non-HIV population vary widely.^57–61^ Based on indexed LV mass in the mUTIMA study,^56^ a standard deviation of 15-20 g/m^2^, and a two-sided α of 0.05, the ASANTE study will have 99-100% power to detect a similar difference (~10 g/m^2^) by HIV serostatus.

The ASANTE study will also investigate the relative contribution of latent tuberculosis infection (LTBI) to these outcomes. Detectable effects of LTBI on primary outcomes based on a sample size of 400 participants are illustrated in **Table 3**, accounting for a range of prevalences of outcomes and exposure given that data on HIV-associated cardiovascular changes in Africa remain sparse. The current evidence estimates LTBI prevalence in East Africa to be as high as 48-49%^40–42^ among persons with and without HIV.

**Table 3.** Minimal detectable odds ratios for outcomes (subclinical coronary and myocardial abnormalities) based on n = 400, 5% type I error, and 80% power.

| Latent tuberculosis infection exposure prevalence<sup>†</sup> | Prevalence of outcome in unexposed group<sup>‡</sup> |  |  |  |
| --- | --- | --- | --- | --- |
|  | 10% | 20% | 30% | 40% |
| 30% | 2.32 | 1.94 | 1.81 | 1.78 |
| 40% | 2.14 | 1.82 | 1.72 | 1.68 |
| 50% | 2.06 | 1.77 | 1.68 | 1.64 |
| 60% | 2.04 | 1.76 | 1.67 | 1.64 |

<sup>†</sup> Latent (LTBI) and active tuberculosis (TB) prevalence is presented as a range based on literature from the general and HIV populations.
<sup>‡</sup> As estimates of structural cardiovascular changes are limited in the region, a broad range is presented based on the available literature.

### Statistical analyses

Separate multivariable regression models adjusted *a priori* for age and sex will be used to estimate the association of each exposure of interest (HIV, cardiometabolic risk factors, LTBI status) with the coronary and myocardial outcomes of interest (**Table 2**). Multivariable logistic regression will be used to model binary outcomes, while multivariable linear regression will be applied to continuous outcomes. We will also assess left ventricular mass as a continuous outcome in multivariable linear regression models adjusted for height and weight to account for indexed LV mass, age, and sex. Non-normality will be addressed using an appropriate transformation of continuous variables. Linear model assumptions will be assessed with residual analysis. Combined models will be run after assessing for multicollinearity between exposures using variance inflation factors. Additionally, an interaction between each exposure and HIV status will be used to determine if the association between each exposure and outcome is modified by HIV serostatus. In models restricted to individuals with HIV, we will also adjust for HIV-specific characteristics (e.g., nadir and current CD4^+^ T-cell count, viral load). A framework for planned primary analyses is shown in **Figure 2**.

**Figure 2.**
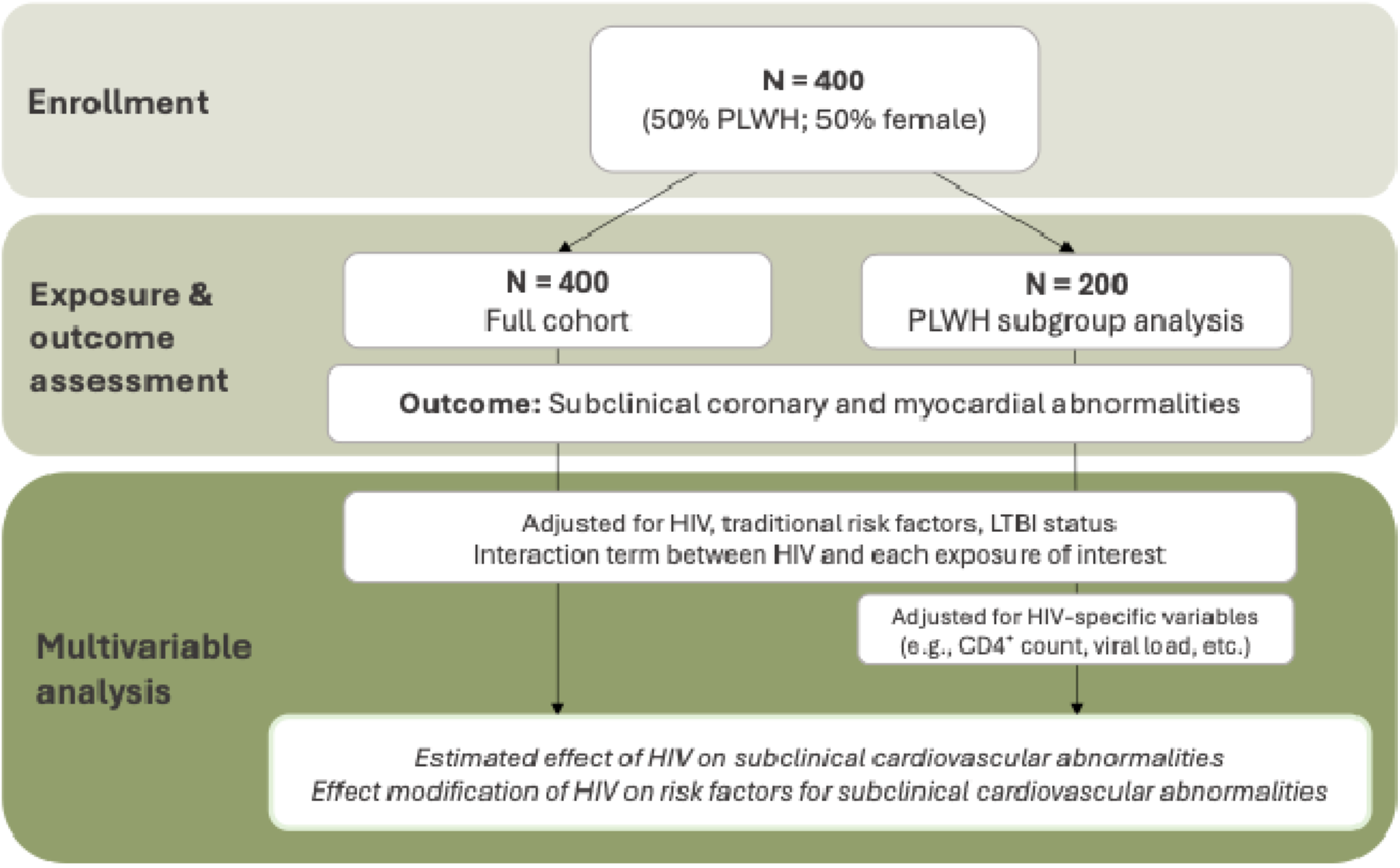
Primary analyses to be performed in the ASANTE study (LTBI, latent tuberculosis infection).

### Study timeline

Preparations for data collection, including protocol development, ethical approval, and staff training, took place from January to December 2023. Data collection commenced in January 2024 and is expected to be completed in December 2027. Data analysis will begin on completion of data collection.

### Limitations and Potential Impact

#### Limitations

This study has limitations associated with all observational research, including potential unmeasured residual confounders and inability to prove causality due to the cross-sectional design. Collecting baseline data, expanding the sample size, and performing longitudinal follow-up assessments will increase power to assess the correlation between changes in variables over time; this will provide additional validity to cross-sectional associations. Further, as this is a single-center study, there is a concern that findings may not be generalizable to other populations; to mitigate this, results from ASANTE can be compared with similar cardiac imaging studies in East Africa, such as the Ugandan Study of HIV Effects on the Myocardium and Atherosclerosis (mUTIMA).^5,23,46,62–64^ The mUTIMA study enrolled 200 participants (50% PLWH), and data from ASANTE can be compared and pooled to examine location-specific risks and disease patterns.

#### Impact

This cross-sectional study aims to investigate the prevalence of and risk factors for early structural cardiovascular abnormalities in HIV infection in an urban and peri-urban population in Nairobi, Kenya. To our knowledge, this is the first study to employ multimodal cardiac imaging in Kenya. Specifically, CCTA has not previously been performed for research purposes in this setting. ASANTE will offer novel insights on prevalence and phenotypes of coronary atherosclerosis in this population, leveraging anatomic imaging that is comparable to the gold standard of invasive angiography. Findings from the sensitive echocardiographic assessments performed in ASANTE can be used for tailored risk stratification in this population. Finally, identifying LTBI as a possible endemic risk factor for early structural cardiovascular disease among PLWH in Kenya may inform management with prophylactic antibiotics. Further, biological samples collected in ASANTE can be leveraged in future biomarker and multi-omic studies to evaluate mechanistic pathways underlying HIV- and latent tuberculosis-associated cardiovascular phenotypes in this population. In combination with biological samples, the phenotypic associations may help to uncover novel biochemical pathways that can be targeted with pharmacologic therapies. Finally, the lessons learned from launching this multimodal cardiac imaging study in Kenya can be translated to similar resource-constrained settings. This reciprocal knowledge exchange can facilitate similar studies defining HIV-associated CVD phenotypes in similar settings, leading to improved representation of developing countries in HIV-associated cardiovascular research.

### Ethics and dissemination

#### Ethical considerations

Approvals have been obtained from the Kenyatta National Hospital-University of Nairobi Ethical Review Committee (KNH-UoN ERC Ref. P611/07/2022) and the University of Washington Institutional Review Board (STUDY00015699) to conduct this study. All participants provide written informed consent following a detailed discussion of study procedures and associated risks and benefits in their preferred language (Kiswahili or English). Any clinically actionable incidental laboratory or imaging findings uncovered during data collection are referred for appropriate management to local medical providers. Eligible participants are assigned a study ID at enrollment under which all data are catalogued; no identifiers are stored with any collected data, thereby ensuring privacy and confidentiality.

Although this is an observational study, one of the study procedures used to assess outcomes, CCTA, confers more than minimal risk due to pre-medications required for optimal image quality, radiation exposure, and iodinated contrast administration. To mitigate these risks, screening criteria exclude participants who have contraindications to beta blocker or nitroglycerin therapy (**Table 1**). Radiation exposure is minimized using ECG-gated tube modulation during image acquisition; further, women of childbearing age must have a negative pregnancy test to be eligible for the study. Screening criteria also exclude participants with an estimated glomerular filtration rate (eGFR) <60 mL/min/1.73 m^2^; although iodinated contrast is safe in more advanced kidney disease, we have adopted a more conservative eGFR cutoff due to limited ability to monitor for contrast-induced nephropathy in this resource-limited setting.

#### Dissemination

Study results will be presented at national and international conferences and submitted for publication in peer-reviewed journals.

## Authors’ contributions

SSS and CTL contributed to the conception and design of the study. SSS, SK, GO, BMA, BG, MM, JK, BC, and AOO contributed to study supervision and data collection. SSS drafted the manuscript. All authors participated in critical revision of the manuscript and approved the final version for publication.

## Funding statement

This work was supported by the Fogarty International Center of the National Institutes of Health under a grant awarded to the Northern Pacific Global Health Fellows Program (#D43TW009345, SSS); the Firland Foundation (#20220003, SSS); the University of Washington Global Cardiovascular Health Program (SSS, CTL); the University of Washington Global Innovation Fund (SSS), and the American Heart Association (#24SCEFIA1258761, SSS). The content is solely the responsibility of the authors and does not represent the official views of any sponsoring agencies.

## Competing interests statement

The authors report no conflicts.

## Data Availability

All data produced in the present work are contained in the manuscript.

## Notes

### Competing Interest Statement

The authors have declared no competing interest.

### Author Declarations

Approvals have been obtained from the Kenyatta National Hospital-University of Nairobi Ethical Review Committee (KNH-UoN ERC Ref. P611/07/2022) and the University of Washington Institutional Review Board (STUDY00015699) to conduct this study.

